# The Impact Of Alcohol Minimum Unit Pricing On People with experience of Homelessness: Qualitative Study

**DOI:** 10.1101/2023.03.31.23287966

**Authors:** Carol Emslie, Elena Dimova, Rosaleen O’Brien, Martin Whiteford, Sarah Johnsen, Robert Rush, Iain D. Smith, Tim Stockwell, Anne Whittaker, Lawrie Elliott

## Abstract

**BACKGROUND:** Alcohol Minimum Unit Pricing (MUP) was introduced in Scotland in May 2018. Existing evidence suggests MUP can reduce alcohol consumption in the general population, but there is little research about its impact on vulnerable groups. This qualitative study aimed to explore experiences of MUP among people with experience of homelessness.

**METHODS:** We conducted qualitative semi-structured interviews with a purposive sample of 46 people with current or recent experience of homelessness who were current drinkers when MUP was introduced. Participants (30 men and 16 women) were aged 21 to 73 years. Interviews focused on views and experiences of MUP. Data were analysed using thematic analysis.

**RESULTS:** People with experience of homelessness were aware of MUP but it was accorded low priority in their hierarchy of concerns. Reported impacts varied. Some participants reduced their drinking, or moved away from drinking strong white cider in line with policy intentions. Others were unaffected because the cost of their preferred drink (usually wine, vodka or beer) did not change substantially. A minority reported increased involvement in begging. Wider personal, relational and social factors also played an important role in participant responses to MUP.

**CONCLUSION:** Our findings suggest that MUP worked as intended for some people with experience of homelessness, while a minority reported negative consequences. Our findings highlight the importance of considering the impact of population level health policies on marginalised groups. Policymakers in Scotland, and elsewhere, need to consider wider contextual factors that affect responses to MUP in people with experience of homelessness. It is important to invest further in secure housing and appropriate support services for people with experience of homelessness who consume alcohol, and implement and evaluate harm reduction initiatives such as managed alcohol programmes.

## INTRODUCTION

The Scottish Government introduced Minimum Unit Pricing (MUP) in 2018 following an extensive legal case involving the global alcohol industry (Carlin, 2022; Katikireddi et al., 2014). There is strong evidence that reducing the affordability of alcohol lowers alcohol consumption and alcohol related harms (World Health Organization. Regional Office for Europe, 2021). The underlying rationale of MUP is therefore to set a minimum price at which alcohol can be sold; it is primarily targeted at inexpensive drinks that are typically consumed by heavier drinkers with the intention of reducing alcohol related harms and mortality (World Health Organization. Regional Office for Europe, 2022). Scotland is one of the first countries to adopt this particular approach (World Health Organization. Regional Office for Europe, 2022). Other pricing policies operate in Canada, Russia, and some former Soviet Union countries. More recently, MUP was introduced in Australia (Northern Territory), Wales and the Republic of Ireland (World Health Organization. Regional Office for Europe, 2022).

The best available evidence for effectiveness comes from modelling and real-world research. Globally, studies suggest that minimum pricing is effective in reducing alcohol-related morbidity, harms and alcohol consumption. Notable examples indicate a reduction in morbidity among poorer communities in the UK and South Africa (Brennan et al., 2021; Gibbs et al., 2021) and a reduction of mortality and alcohol harms in Canada (Stockwell et al., 2020; Zhao et al., 2013). Minimum pricing is also associated with reductions in hospital admissions in Canada, in particular among lower income families (Zhao & Stockwell, 2017) and reduced alcohol related admissions to intensive care units in Australia (Secombe et al., 2021). It is also associated with greater reduction in consumption among harmful drinkers compared with moderate drinkers in Australia (Jiang et al., 2020). Research also suggests that MUP reduces the consumption of inexpensive high alcohol beverages in Australia (Taylor et al., 2021) and the general consumption of alcohol, in particular among households that purchase greater quantities of alcohol, in Wales and Scotland (Anderson et al., 2021). General reduction in alcohol consumption was also observed in Russia after the introduction of MUP (Neufeld et al., 2020; World Health Organization. Regional Office for Europe, 2019).

MUP has been extensively evaluated in Scotland, more so than in any other country (Beeston et al., 2020; Carlin, 2022; World Health Organization. Regional Office for Europe, 2022). The findings of these evaluations broadly reflect those of existing studies: for example a reduction in alcohol consumption in the general population (Giles et al., 2021) in particular among households that purchase greater amounts of alcohol and those with lower incomes (O’Donnell et al., 2019). Alcohol specific deaths fell by around 10% in Scotland in 2019, before rising again during the COVID pandemic (Angus, 2020; Holmes, 2023). Some research has focused on the impact and experiences of MUP among marginalised groups such as harmful and dependent drinkers (Holmes et al., 2022). Concerns that dependent drinkers would substitute illicit or non-beverage alcohol (i.e. surrogate alcohol not intended for consumption such as mouthwash or hand sanitizer) or other substances, or steal if they could not afford their drink of choice were not borne out in practice (Holmes et al., 2022). However, Holmes et al (2022) found no clear evidence that MUP led to a reduction in the consumption of alcohol and some evidence of financial strain.

None of the studies described above explore the lived experience of MUP among people experiencing homelessness. Recent figures suggest that 14,250 households in Scotland experienced homelessness on any given night in 2019, including rough sleeping, sofa surfing, staying in hostels, refuges or unsuitable forms of temporary accommodation (Watts et al., 2021). These highly marginalized populations may be disproportionately affected by pricing interventions given their limited income and consumptions of strong and cheap alcohol (Goodall, 2011; McGill et al., 2016).

### Aim

Our study used a qualitative approach to explore the impact of MUP in Scotland on people experiencing homelessness. It builds on previous research which explores strategies adopted among this population when alcohol becomes unaffordable (Erickson et al., 2018).

## METHODS

This qualitative study used individual interviews to elicit in-depth accounts from people who reported a history of homelessness and of drinking alcohol before the implementation of minimum unit pricing (MUP) in Scotland on 1^st^ May 2018. Following Bramley (2017), experience of homelessness included rough sleeping, staying in places not intended for habitation (e.g. cars, tents, sheds), and/or living in temporary or insecure accommodation (e.g. hostels, refuges, sofa surfing). The study was conducted in Glasgow, the largest city in Scotland, as its population demographics and alcohol consumption patterns represent a particular challenge. Compared with the rest of Scotland, it has the highest levels of deprivation (Scottish Government, 2021), the highest number of premature deaths from alcohol (Schofield et al., 2021, 2021), and the highest number of people experiencing homelessness prior to the COVID-19 pandemic (National Statistics, 2020). All procedures were performed in compliance with relevant laws and institutional guidelines. As our study involved human participants, ethical approval was required and was granted by the Nursing Department Research Ethics Committee at Glasgow Caledonian University (HLSNCH18029) in May 2019, followed by an amendment to conduct remote interviews due to COVID-19 restrictions in July 2020.

We set up a study steering group to advise on the academic, ethical and policy aspects of our research. This included academics with topic expertise, health professionals and representatives from charities working with people with experience of homelessness. We also convened a stakeholder group which included those with lived experience of homelessness, representatives from the study partner organisation (Homeless Network Scotland) and other third sector organisations, police representatives, policy makers and academics. Both groups advised on methodological approaches, commented on interpretation of findings and helped formulate policy recommendations.

The study also involved interviews with stakeholders working with people experiencing homelessness to explore their views on MUP; these findings are published elsewhere (Dimova et al., 2023).

### Recruitment and participants

Recruitment took place over two phases: October 2019 to January 2020 (before the COVID-19 pandemic) and August to October 2020 (during the COVID-19 pandemic). We used a purposive sampling strategy to ensure we included a diverse sample of participants by gender, age and housing status. To be included in the study, participants had to be aged 18 years or over, have experience of homelessness, have drunk alcohol before the implementation of minimum unit pricing on 1 May 2018, be able to converse in English and have the ability to provide informed consent.

In the first phase, a qualitative researcher with previous experience with homeless populations (MW) worked closely with the study partners, The Homeless Network, and other community, faith and voluntary sector organisations to facilitate site visits and face-to-face recruitment. Participants were recruited from a day centre for people with experience of homelessness, an open access support hub, a women-only accommodation service and a crisis residential unit for people with complex needs and a history of housing vulnerability or rough sleeping. In the second phase, participants were identified by gatekeepers within support services who approached potential participants, distributed participant information sheets and answered questions. In all cases, informed consent was obtained by the researcher. In total, we interviewed 46 people with current or previous experiences of homelessness. Of these, 18 also reported involvement in street drinking (i.e. regularly drinking outdoors in public places because they did not have access to domestic space where alcohol consumption is permitted and/or were unwelcome or could not afford to drink in pubs: Johnsen et al 2010). The demographic characteristics of interviewees are shown in Table 1

**Table 1.** Participant characteristics (n=46)

|  |  |
| --- | --- |
| **Age** |  |
| Mean | 47 years |
| Range | 21-73 years |
| **Gender** |  |
| Men | 30 |
| Women | 16 |
| **Race/ethnicity** |  |
| White Scottish | 45 |
| White Other | 1 |
| **Housing status** |  |
| Supported Accommodation | 16 |
| Specialist Alcohol Detox Unit | 11 |
| Hostel | 7 |
| Emergency Accommodation (during the COVID-19 pandemic) | 4 |
| Private Accommodation | 3 |
| Other (social housing, sheltered housing, sofa surfing, rough sleeping, Housing First programme) | 5 |
| **Drinking status (AUDIT-C)\*** |  |
| Possible Dependence (at the time of interview) | 21 |
| Increasing Risk | 5 |
| Low Risk | 9 |
| **Use of illegal drugs** |  |
| Current or recent use of illegal drugs | 9 |
| Former use of illegal drugs | 10 |
| Use of prescribed medication (including methadone) | 22 |

\*The AUDIT-C was not administered to 11 participants who were in a detox facility at the time of interview.

## Data collection

In phase one, face-to-face interviews were conducted with 25 respondents. All were individual interviews apart from two paired interviews, undertaken at the request of women in the female-only accommodation service. Phase two interviews (n=21) were conducted by telephone during the COVID-19 pandemic. Gatekeepers within support services facilitated access to phase two interviewees by distributing information sheets and inviting eligible service users to consider contributing. These interviews took place in a private settings either in the service (e.g. an office) or participants’ accommodation. All respondents were asked to complete a socio-demographics questionnaire and the AUDIT-C (a short version of the Alcohol Use Disorders Identification Test, a validated alcohol harm assessment tool which indicates higher risk drinking or possible alcohol dependence: (Bush et al., 1998; Saunders et al., 1993) with the exception of those in a detoxification service where administering AUDIT-C was not appropriate.

All interviews were audio recorded, with the participants’ permission, transcribed verbatim by a professional transcriber and anonymised. Each participant was given an ID number not known to anyone outside the research study. The topic guide focused on awareness of minimum unit pricing, and the impact on drinking and alcohol affordability, health and wellbeing and engagement with services. The interviewer tried, wherever possible, to draw out and differentiate between the impacts of MUP vis-a-vis other policies and contextual factors. All participants were given a £15 high street voucher to thank them for their help.

### Data analysis

Data were analysed using thematic analysis (Braun & Clarke, 2006) and facilitated by NVivo v12 software. After data familiarization and discussions within the research team, an initial descriptive coding frame was developed based on the topic guide and a sample of early transcripts. Initial descriptive data summaries focused on: 1) Current alcohol and drug use; 2) Awareness and perceptions of MUP; 3) Strategies to buy alcohol; 4) Effects of MUP. Next, RO independently coded the complete dataset, mapped each theme ensuring that all relevant data were captured, developed higher level codes, identified additional concepts and themes, and refined the analytic account of the data. Emerging findings were discussed at weekly research meetings – with all authors contributing to analysis - and with our steering and stakeholder groups. These discussions generated further questions explored by RO such as identifying and comparing subgroups of participants that were of interest analytically (e.g. reduced drinking due to MUP v no change), separating respondents’ accounts of their own behaviour from accounts of other people’s behaviour, and trying to differentiate the impact of MUP from COVID-19 and other significant events in people’s lives. At this stage, we sought to organize our findings around the objectives of MUP, principally awareness of the policy, and reported impacts such as intended outcomes and unintended consequences.

## FINDINGS

### A) Background: Awareness and priority accorded to alcohol minimum unit pricing

Most respondents reported they were aware of the introduction of alcohol minimum unit pricing in Scotland. They were particularly aware of the steep rise in the cost of strong white ciders. While some recognized and supported the intention to reduce the consumption of cheap, strong alcohol [‘*that cider was killing people, mate’ #39*], and hoped that it might encourage people to seek help earlier [*it’s gonna be beneficial - maybe seek help sooner than later #48]*, there was some concern that MUP would disproportionally affect poorer people, particularly those dependent on alcohol. It was notable in respondents’ accounts how they drew on different models of addiction (e.g. disease model of addiction, addiction as self medication, addiction as a social justice issue) to criticize this policy. For example, a dominant narrative in the interviews about the powerlessness of dependent drinkers to stop drinking was deployed to suggest that this increase in price was both ineffective and unfair for this group [‘*if you’re an alcoholic and you crave that drink, you’re gonna get it one way or the other’ #56*]. Several other respondents were concerned that MUP, in isolation, would not address the inequalities perceived to underlie and exacerbate alcohol problems in people with experience of homelessness:-

> *I think if we had*.. *a better society where*.. *what we have is distributed a bit more fairly, I think people wouldn’t feel the need to self-medicate*.. *quite so much*
>
> *(#49, female)*
>
> *Society’s got*.. *a horrible view of people with addictions. The Government are all rich*.. *they’ve never had to live how we had to live*.. *Help them (people with experience of homelessness) with whatever it is that causes them to drink and causes them to do all that stuff. Then you’ll find they might not have to put fucking price rises on things ‘cause people are not gonna abuse it (substances) as much*.
>
> *(#35, male)*

Despite awareness of the introduction of minimum unit pricing, most respondents did not perceive it to be particularly salient to their daily lives. Respondents freely discussed their current struggles, experiences of drinking, health problems and history of homelessness but often needed repeated encouragement to discuss MUP. This appeared to indicate that it had low priority in their hierarchy of concerns given the difficulties they faced on a day -to day basis, including the impact of welfare reforms, challenges accessing accommodation, welfare benefits and treatment services, and coping with mental and physical health problems:-

> *No, we don’t discuss it (MUP) at all (in the hostel). We might moan about like, ‘fuckin’ hell,’ know what I mean, ‘like it’s (price) went up*…*’ But we don’t moan about minimum price per unit, that’s not a conversation*.. *people have*.
>
> *(#41, male)*

### B) Reported Impact Of MUP

Accounts suggested that MUP had a range of impacts on drinking practices and related behaviours. Some reported that it did not affect them at all, some reported that they had reduced their drinking or moved away from consuming strong white cider (in line with policy intentions), while others discussed possible unintended consequences of the policy.

#### No impact on alcohol consumption

Some respondents reported that their drinking was unaffected because the cost of their preferred beverage (usually wine, vodka or beer) did not change substantially after the introduction of MUP. This group often contrasted the lack of change in price of their preferred drink with the huge rise in the cost of strong white cider:-

> *Cause it was only really wine that I would buy, so…It didn’t really impact on me, but*.. *the cider…*
>
> (#18, female)
>
> *I drink (cheap vodka brand)*.. *I didn’t see the change (in price). But I know people that I’ve talked to in the street that used to drink these bottles, like litres of (strong white cider) I wasn’t affected by it … it’s just not my drink of choice*.
>
> (#2, male)
>
> *I just drink beer, and it didn’t really affect the price of beer that much, so I’m not really a spirits drinker or a cheap kinda cider (drinker)*.
>
> (#41, male)

#### Reduced alcohol consumption (intended consequence of MUP)

A small group of respondents – all women – reported that they had reduced their drinking, at least partly because of the increase in price, after the introduction of MUP (that is, responded in line with policy intentions). All of these women discussed how serious health concerns (their own or a relative’s) influenced this decision, alongside price. For example, one woman was taking medication for a serious health condition, another had recently been discharged from hospital and a third discussed her husband’s death due to alcohol. The first excerpt below (#9) provides a good illustration of how a number of different health and social factors could combine to result in behavioral change (in this case, a population level measure such as MUP, alongside personal health concerns and the desire to see her children):-

> *I’ve had a major problem with alcohol*..*but I’m just managing to sorta kick (it) because of the increase in prices… (With) the medication I’m on*.. *that makes me feel sick anyway*… *I’ve got four kids*.. *I want them to know who I am without that side of the drink*… *So*.. *I have cut down*.
>
> *(#9, female. Reported reducing drinking from 3 litres of cider daily to vodka once or twice a week*.)
>
> *It’s dead dear [expensive] for drink now (after MUP)*.. *I changed my drinking. I used to drink*.. *superlagers – I drank 8 of the cans. I wanted to come off the drink a lot (because)*.. *it’s dead dear*.
>
> *(#8 female. Reported reducing drinking from 8 cans of superlager or a litre of vodka daily to not drinking any alcohol*.)
>
> *Aye, it’s (drinking) went down*.. *it’s down a bit, you know? ‘Cause you’ve got to watch, it’s getting dearer*.
>
> *(#28 female. Reported reducing drinking from 1 – 2 bottles of vodka to 4-5 cans of alcoholic energy drinks daily. Recently left hospital)*.

#### Move away from consumption of cheap strong white cider (intended consequences of MUP)

More commonly, respondents reported switching beverages, usually from strong white cider (which had increased dramatically in price from £4.50 to £11.25 per 3 litre bottle - 22.5 units) to spirits (often vodka – priced around £13 for 26 units). This move away from strong, cheap cider was an intended aim of MUP. However, some respondents saw switching drinks as a way of ‘getting round’ MUP (*It never put me off. All it did was make me change my drink…#52*). Given that a bottle of vodka was now priced similarly to a 3 litre bottle of white cider, vodka was perceived to be good value (#52) as it lasted longer (using a mixer) and was ‘*the quickest way*..*to get drunk’* (#13) and ‘*get a buzz straight away’* (#14).

> *Oh, why are you buying (strong white cider brand), mate? You can get a bottle of vodka*.*’ ‘Well you see I was saying that because it was round about the same price. And the vodka gets you drunk*.
>
> *(#43, female)*
>
> *I went onto the vodka rather than getting [cider], ‘cause I was basically budgeting*.. *I thought ‘if I get x amount of cans now at that price, then it’s gonna cost a lot more, so I’ll just get a bottle (of vodka)’*
>
> *(#45, female)*
>
> *I don’t know if it affected the amount of alcohol I’d take, but it definitely affected what type of alcohol I was taking*.
>
> *(#51, male: reported switching from strong white cider to fortified wine)*

A few respondents who had switched to vodka worried that this had a (more) detrimental effect on their health. For example, despite reducing her drinking after MUP (see above), one woman (#9) was concerned that vodka was more damaging than strong cider. She also described finding it easier to ‘control’ her intake when drinking cider. Others were concerned about the impact of spirits on their stomach. However, as the extract below (#52) suggests, it was difficult for some respondents to separate the impact of drinking cider for years (6 litres of strong cider daily) from the switch to spirits:-

> *To be honest, I feel as if I would prefer to drink the cider than*.. *vodka because it’s the effects - I know I’m doing myself damage. Cider*.. *it’s like a juice, it’s easily controlled. Vodka*..*has more effect on*.. *your mental and physical state*.. *If the prices went back to the way it was (before MUP) - it’s not just myself - everybody would literally go back to your wee (strong white cider brand), eh?*
>
> *(#9, female)*
>
> *When I went onto spirits (from cider)…it was actually worse for me than it was beneficial. I don’t know if it was because of the cider, but your stomach used to get quite bad from it*.
>
> *(#52, male)*

#### Unintended consequences of minimum unit pricing

Respondents described a range of ways they obtained alcohol when money was limited prior to the introduction of MUP, most commonly pooling resources (money or alcohol), taking turns buying alcohol with friends (alternating depending on receipt of welfare benefits) and borrowing money. Other strategies included being paid for casual jobs, being given a few cans of lager for buying alcohol for others, going without food or cigarettes to prioritise money for alcohol, pawning possessions, switching to cheaper alcohol, spending less on alcohol, begging or stealing alcohol (or asking others to steal alcohol for them). When respondents reported begging to raise money for alcohol, they often stressed that this only occurred at particularly difficult times: ‘*just when I needed it’* (#14), *‘when I’m desperate’* (#37) or when ‘*I’ve been on the DTs* (i.e. delirium tremens)’ (#4).

Respondents continued to use these strategies after the introduction of MUP. Linked to the dominant narrative discussed above, there was a common perception that **other** people would do whatever was necessary to obtain alcohol, even when the price rose. However, fewer respondents reported that their **own** behaviours were affected by MUP. One woman described stealing from her family to finance her drinking (taking the ‘*odd pound here and there’*). However, as the price of cider increased after MUP, stealing from her family became more noticeable, so she described getting others to steal cider for her, or stealing alcohol herself, and then finally switching to buying spirits for economic reasons and fear of being caught:-

> *I did get in a bit of bother with it because of the pricing. I would get somebody to steal it… or there is a couple of times I’ve tried myself. But*.. *(you don’t) want to get caught …(and) what’s the point in (asking) somebody*.. *to steal a bottle of cider when to be honest you’re cheaper and better off buying your spirits?*
>
> *(#9, woman)*

A few respondents explicitly linked MUP to changes in their own begging behaviour (i.e. begging more frequently or for longer periods of time to get the money required to buy alcohol). People who beg are often victims of crime and antisocial behavior (as noted by #17 in the extract below, as well as by other respondents:-getting ‘*mugged*.. *battered*.. *getting (money) cups lifted’* #3, ‘*they’ll run up and grab your cup and bolt’* #4). Changes in begging behavior linked to MUP are therefore important as increased time on the street heightens the widely documented risk of violence or theft (Johnsen 2010):-

> *(MUP) didn’t have to go that high*.. *A can of (strong cider), it was only £1, Now (after MUP), it’s £2…More begging to get more money*.
>
> *(#14, male)*
>
> *Before this price came in (MUP), they were £1*..*then they made it £2 a can which I was raging at*.. *So it really affected me cause I was*..*out begging for money*.. *You’ve got £2 and you think ‘yes! That’s 2 cans’, and then you remember, ‘oh no, it’s 1 can’*.
>
> *(#35, male)*
>
> *You’ll sit and beg for an extra hour or two, to make that extra money (after MUP)*.. *I’ve been in that situation, I had to get maybe an extra pound for four cans, and I’m sitting, and nothing happening, nothing happening, and*.. *then somebody just drops a pound in your cup, and you’re like, right, straight off to the offies,(off-licence). (Later)*.. *But the experience of being homeless, it really hurt me deep down, you know? I hit rock bottom a couple of days ago, I was drinking a can of lager, somebody stole it off me. Four people attacked me. I got a black eye*.
>
> *(#17, male)*

Another common perception was that minimum unit pricing would push (other) people who had experienced homelessness to ‘*turn to drugs*.. *find their buzz or their fix elsewhere*’ (#45) particularly given the increased availability and cheap price of ‘street’ benzodiazepines in Glasgow at the time (see McAuley et al., 2022). For example, one man (#2) suggested that after the rise in the price of strong white cider, it made economic sense to switch to ‘street’ valium, while a street drinker (#3) sadly referred to the deaths of a number of his friends which he perceived to be linked to this switch. (It should be noted that public health officials had recently suggested that the sale of ‘street’ drugs sold as valium was linked to a very high number of fatal and non-fatal related overdoses in Glasgow, including among people with experience of homelessness: Glasgow City Council, 2019):-

> *I think there’s lot o’ drugs moved in…If they’ve got £10*.. *well, they were gonna buy three bottles of (strong white cider before MUP). Now*.. *they’re going, “Right, well fuck it, you can get 25 (street) Valium*.*”*.. *and just split the (street) Valium between them …getting them to that stage that they want to be after a bottle of (strong white cider)*.
>
> *(#2, male)*
>
> *But since the drink price has went up*,..*people are turning to other things*.. *See just in the last two month? I’ve lost four of my pals… And they’ve all passed away with taking (street) Valium. These are guys that were on the drink … they can’t afford to drink cider or a bottle … Because drink has got dear, that’s how you’ve got so many deaths now, because they’re all turning to (street) Valium. Their bodies has been used to drink*.
>
> *(#3, male)*

There was no evidence that respondents who had not previously used illicit drugs started to use them after the introduction of MUP. However, a few respondents who described using both alcohol and other drugs before the introduction of MUP suggested that MUP may have influenced the balance of their alcohol consumption compared to other substances. Perhaps surprisingly, two women (both street drinkers in supported accommodation who took part in a paired interview) suggested that MUP may have led to a reduction in the amount of cocaine they bought:-

> *Do you know why it (MUP) didn’t change anything for any of us really? Cause, right, see if I had to get a bit of coke (cocaine) when you’re drinking, right? The bit of coke depends on how much drink you buy. So if you’re buying the drink, whatever’s left is going on coke anyway. So it’s not as if it (alcohol) would’ve changed. Might change how much coke you’re (buying)*.
>
> *(#29 and #30)*

In contrast, two respondents linked MUP to their increased use of illicit drugs. One woman (#48) described how the increase in the price of strong cider resulted in a reduction in her alcohol consumption and an increase in consumption of ‘street’ benzodiazepines. However, she still supported the introduction of MUP as she felt that it would encourage some people to seek help more quickly:-

> *It (cider) was just so unbelievably cheap but I’m so glad they put the price up to be honest with you…But I couldn’t afford to drink as much as I did*.. *Unfortunately at the same time (as) drinking, I was taking drugs as well. So it (MUP) kinda affected things*.. *(I was taking) more drugs*.. *Everybody was the same, the street Valium and things like that … Obviously it helped me not drink as much*.. *You could get like 25 tablets (street Valium) and that would last me*.. *like three days as well as like the alcohol. But yeah … money-wise… taking the tablets and less drink kinda evened me out*.
>
> (#48 female)

One man (#44) described selling drugs before MUP and using the profit to buy alcohol. He described how the increase in the price of alcohol meant he needed to sell more drugs to make more money ‘so that made me more of a criminal’. However, he had been diagnosed with a chronic disease in 2018, which made it difficult to separate the impact of MUP from health and mobility issues. He described how this condition led to him drinking less alcohol and self-medicating with cannabis. Again, the relationship between increased cost and decreased use was not clear-cut. He described how in the past he was also taking cocaine ‘that cost more than booze’, and so reflected that the price of alcohol was not the only factor affecting his behaviour:-

> *I was a wee bit of a criminal, and I sold a bit of drugs*.. *and tried to get by that way*... *So I could*..*make some money, pay the bills that I had to pay, and then I’d have profit to buy my booze. (MUP)*.. *made me sell more drugs, so that made me more of a criminal*.
>
> ***Interviewer: So do you think there is a connection between the change in price, and your criminality, do you think? Or is that too simplistic to say that?***
>
> *It’s too simplistic to say that*.. *because*.. *I’ve got (chronic disease) mate, so that changed a lot. So the unit pricing and the (chronic disease) mix together…*.*I can’t walk to go to the toilet*.. *So I try to stay away from the drink. (LATER)*.. *My life was chaotic, and I was taking whatever to get mad with it, and it didn’t matter the cost, cause I was taking coke (cocaine) for a while, that cost more than booze*.
>
> *(#44, male)*

Finally, it was notable that none of our respondents with experience of homelessness made any reference to drinking non-beverage alcohol (surrogate alcohol not intended for human consumption).

## DISCUSSION

This is the first qualitative study to explore the impact of alcohol minimum unit pricing among people who have experience of homelessness. We found that most respondents were aware of MUP and discussed the steep rise in the cost of some types of alcohol. The extent and nature of their responses to MUP varied. In line with policy intentions, some respondents reported reducing their alcohol consumption because of MUP or moving away from drinking strong white cider. Some were unaffected because the price of their preferred drink did not change. Unintended consequences included a possible increase in involvement in begging to enable respondents to maintain their level of alcohol consumption. In these qualitative accounts, there was no evidence that MUP had resulted in substitution to non-beverage alcohol, and little suggestion that it had directly resulted in substitution to illicit drugs. Overall, most respondents perceived MUP to be relatively low in their hierarchy of concerns, given the complexities and hardships of homelessness.

Reduction in alcohol use, in particular the consumption of inexpensive drinks such as strong white cider, has been observed in quantitative research from Australia (Taylor et al., 2021), Canada (Stockwell et al., 2012) and Scotland where a reduction in alcohol consumption was found among lower income households (O’Donnell et al., 2019). It was notable in our study that all the respondents who reported reducing their drinking were women. Further research is required to ascertain whether there is a gender difference in these impacts. It could be that women were already motivated by health and family concerns to change their drinking behaviours, and that MUP provided the final nudge, or it could be that women were more willing to report reducing their alcohol consumption (see, for example, Lyons et al (2014) where female drinkers were much more likely than male drinkers to describe stopping drinking when they realised they had consumed too much alcohol and were approaching their ‘limit’).

While no other qualitative study of MUP has focused on people with experience of homelessness, there has been some research with other marginalized groups. Holmes et al (2022) explored the impact of MUP on harmful and dependent drinkers in Scotland and reported some evidence of reduction in alcohol consumption and switching from strong cider to vodka. Coping strategies were based on previous experiences when money was tight including cutting back on food, increasing borrowing and using food banks. There was no clear evidence of other negative consequences. In contrast, our study found that some respondents linked the introduction of MUP to increased involvement in begging which represents a heightened risk of exposure to violence and theft. It was notable that none of our respondents with experience of homelessness made any reference to drinking non-beverage alcohol, which contrasts with findings from homeless populations in other countries (Erickson et al., 2018; Westenberg et al., 2021). Given the qualitative nature of our research, it was not possible to quantify the most commonly used coping strategies to secure alcohol when respondents could not afford to buy it (see, for example, Erickson et al., 2018).

Particular strengths of our research include our sampling strategy through which we recruited men and women from a wide range of settings, reflecting various types of homelessness. However, all but one person defined themselves as ‘White Scottish’, so the voices of ethnic minorities may be under-represented in our study. One analytical issue was the difficulty in isolating the impact of MUP from other concurring factors that undoubtably affected alcohol use such as poor physical health, the availability of inexpensive illegal drugs such as ‘street’ benzodiazepines, family relationships and the day-to-day difficulties of being homeless. This said, there are examples in our analysis where the impact of MUP is clearly described in participants’ accounts and these are presented above. Finally, given the qualitative nature of our research and the cross-sectional design, quantitative assessments of the change in key behaviours, such as alcohol consumption, were not possible.

Our findings have relevance for policymakers in Scotland, given the ‘sunset clause’ means that the Scottish Parliament must vote to decide if MUP will continue (Beeston et al., 2020), and in other countries that have introduced, or are considering, MUP. In line with policy intentions, some respondents reduced their drinking because of MUP. Others were unaffected because the price of their preferred drink did not change. The level of the unit price of alcohol is salient for both responses. It is important that the unit price reflects economic pressures such as inflation and that more expensive alcohol (wine, vodka and beer) is captured in the policy. It is also important to continue to monitor the impact of MUP on people who have experienced homelessness if the level of the unit price of alcohol is increased. Stakeholders working with homeless populations in Scotland have expressed concern that an increase in the level of MUP without the provision of additional support could increase unintended consequences for this group (Dimova et al., 2023). This links to broader concerns about the impact of population measures on marginalised groups. As Rehm & Probst (2018) have argued “it is not appropriate simply to extrapolate from risks associated with alcohol use in higher-income populations to address lower income populations … Alcohol policy measures not only need to be effective and cost-effective but also need to contribute to a reduction of health inequalities” (p2).

Our findings also have implications for further investment in the provision of appropriate support and secure housing for this marginalised group. There is greater co-morbidity among people who have experience of homeless compared with the general population (Fazel et al., 2014; Palepu et al., 2013; Zeitler et al., 2020) and this should be reflected in the way services work with people who have experience of homelessness (Dimova et al., 2023). Conversations about MUP may stimulate referrals to a range of services highlighting the importance of a national framework for harm reduction and funding mechanisms to allow services to collaborate more effectively.

Similarly, secure housing should be prioritised. Service providers in Scotland, when commenting on the need for additional support for people with experience of homelessness alongside the introduction of MUP, emphasised the lack of stable housing for people at risk of a whole range of harms (Dimova et al., 2023). Housing First was frequently mentioned as an excellent solution because it offers settled housing and holistic support and has been shown to be effective in Scotland as in other countries (Johnsen et al, forthcoming; Woodhall-Melnik et al 2016).

Our findings also suggest the need to implement and evaluate innovative harm reduction initiatives such as managed alcohol programmes (MAPs) which emerging evidence suggest could be promising. MAPs provide measured, regular doses of alcohol and provide a route into harm reduction otherwise not open to those who experience ‘problem’ alcohol use and homelessness (Pauly et al., 2019, 2021). Erickson et al (2018) found that people with housing instability and alcohol dependence who used managed alcohol programmes in Canada were less likely to use illicit drugs and more likely to access treatment when they could not afford to buy alcohol. The need for alcohol harm reduction and managed alcohol programmes for people experiencing homelessness in Scotland has been established (Carver et al 2021, Parkes et al 2021). The first pilot managed alcohol programme in Scotland opened in 2021 but has yet to be fully evaluated.

## Conclusions

Our findings demonstrate a range of responses to MUP from people experiencing homelessness. These include a reduction of alcohol use as intended by the policy. Although most respondents perceived MUP to be low in their hierarchy of concerns, there was evidence of possible unintended consequences such as increased involvement in begging that may have resulted in additional hardship for a minority. Policymakers need to consider the potential impacts on marginalised groups when designing population-wide policies such as MUP. For people who have experienced homelessness, this means investing in secure housing and support services alongside MUP, and implementing and evaluating harm reduction services such as managed alcohol programmes.

## Data Availability

These qualitative data are not publicly available.

## ACKNOWLEDGEMENTS

We would like to acknowledge the important contribution of our partners Homeless Network Scotland, particularly Peter Anderson, as well as Heather Strachan, Rowena Smith, our steering and stakeholder groups, partner organizations who assisted with recruitment and, most of all, the participants.

## References

Anderson, P., O’Donnell, A., Kaner, E., Llopis, E. J., Manthey, J., & Rehm, J. (2021). Impact of minimum unit pricing on alcohol purchases in Scotland and Wales: Controlled interrupted time series analyses. The Lancet Public Health, 6(8), e557–e565.

Angus, C. (2020). Alcohol Deaths and Minimum Unit Pricing. https://www.alcohol-focus-scotland.org.uk/news/alcohol-deaths-and-minimum-unit-pricing/

Beeston, C., Robinson, M., Giles, L., Dickie, E., Ford, J., MacPherson, M., McAdams, R., Mellor, R., Shipton, D., & Craig, N. (2020). Evaluation of minimum unit pricing of alcohol: A mixed method natural experiment in Scotland. International Journal of Environmental Research and Public Health, 17(10), 3394.

Bramley, G. (2017). Homelessness Projections: Core homelessness in Great Britain, Summary Report. Crisis.

Braun, V., & Clarke, V. (2006). Using thematic analysis in psychology. Qualitative Research in Psychology, 3(2), 77–101. https://doi.org/10.1191/1478088706qp063oa

Brennan, A., Angus, C., Pryce, R., Buykx, P., Henney, M., Gillespie, D., Holmes, J., & Meier, P. S. (2021). Potential effects of minimum unit pricing at local authority level on alcohol-attributed harms in North West and North East England: A modelling study. Public Health Research, 9(4).

Bush, K., Kivlahan, D. R., McDonell, M. B., Fihn, S. D., Bradley, K. A., & Project (Acquip, A. C. Q. I. (1998). The AUDIT alcohol consumption questions (AUDIT-C): An effective brief screening test for problem drinking. Archives of Internal Medicine, 158(16), 1789–1795.

Carlin, E. (2022). Regulating the Legal: Minimum Unit Pricing of Alcohol in Scotland. In Ilana Crome, David Nutt and Alex Stevens, Drug Science and British Drug Policy: Critical Analysis of the Misuse of Drugs Act 1971. Waterside Press.

Dimova, E. D., Strachan, H., Johnsen, S., Emslie, C., Whiteford, M., Rush, R., Smith, I., Stockwell, T., Whittaker, A., & Elliott, L. (2023). Alcohol minimum unit pricing and people experiencing homelessness: A qualitative study of stakeholders’ perspectives and experiences. Drug and Alcohol Review, 42(1), 81–93.

Erickson, R. A., Stockwell, T., Pauly, B., Chow, C., Roemer, A., Zhao, J., Vallance, K., & Wettlaufer, A. (2018). How do people with homelessness and alcohol dependence cope when alcohol is unaffordable? A comparison of residents of Canadian managed alcohol programs and locally recruited controls. Drug and Alcohol Review, 37, S174–S183.

Fazel, S., Geddes, J. R., & Kushel, M. (2014). The health of homeless people in high-income countries: Descriptive epidemiology, health consequences, and clinical and policy recommendations. The Lancet, 384(9953), 1529–1540.

Gibbs, N., Angus, C., Dixon, S., Parry, C., & Meier, P. (2021). Effects of minimum unit pricing for alcohol in South Africa across different drinker groups and wealth quintiles: A modelling study. BMJ Open, 11(8), e052879.

Giles, L., Richardson, E., & Beeston, C. (2021). Using alcohol retail sales data to estimate population alcohol consumption in Scotland: An update of previously published estimates. Edinburgh: Public Health Scotland.

Glasgow City Council,. (2019). Drugs Deaths Warning Over Lethal Street Valium [Reference]. 1619. https://www.glasgow.gov.uk/article/23805/Drugs-Deaths-Warning-Over-Lethal-Street-Valium

Goodall, T. (2011). White Cider and Street Drinkers. Recommendations to Reduce Harm. Alcohol Concern. https://rebuildingshatteredlives.org/wp-content/uploads/2013/01/White_Cider_and_Street_Drinkers_report_April_2011.pdf

Holmes, J. (2023). Is minimum unit pricing for alcohol having the intended effects on alcohol consumption in Scotland? Addiction.

Holmes, J., Angus, C., Boyd, J., Buykx, P., Brennan, A., Gardiner, K., Hernandez Alava, M., Hughes, J., Johnston, A., & Little, S. (2022). Evaluating the impact of Minimum Unit Pricing in Scotland on people who are drinking at harmful levels.

Jiang, H., Livingston, M., Room, R., Callinan, S., Marzan, M., Brennan, A., & Doran, C. (2020). Modelling the effects of alcohol pricing policies on alcohol consumption in subpopulations in Australia. Addiction, 115(6), 1038–1049.

Katikireddi, S. V., Hilton, S., Bonell, C., & Bond, L. (2014). Understanding the development of minimum unit pricing of alcohol in Scotland: A qualitative study of the policy process. Plos One, 9(3), e91185.

Lyons, A. C., Emslie, C., & Hunt, K. (2014). Staying ‘in the zone’ but not passing the ‘point of no return’: Embodiment, gender and drinking in mid-life. Sociology of Health & Illness, 36(2), 264–277. https://doi.org/10.1111/1467-9566.12103

McAuley, A., Matheson, C., & Robertson, J. R. (2022). From the clinic to the street: The changing role of benzodiazepines in the Scottish overdose epidemic. International Journal of Drug Policy, 100, 103512.

McGill, E., Marks, D., Sumpter, C., & Egan, M. (2016). Consequences of removing cheap, super-strength beer and cider: A qualitative study of a UK local alcohol availability intervention. BMJ Open, 6(9), e010759.

National Statistics,. (2020). Homelessness in Scotland: 2019-2020. https://www.gov.scot/publications/homelessness-scotland-2019-2020/documents/

Neufeld, M., Bunova, A., Gornyi, B., Ferreira-Borges, C., Gerber, A., Khaltourina, D., Yurasova, E., & Rehm, J. (2020). Russia’s National Concept to Reduce Alcohol Abuse and Alcohol-Dependence in the Population 2010–2020: Which Policy Targets Have Been Achieved? International Journal of Environmental Research and Public Health, 17(21), 8270.

O’Donnell, A., Anderson, P., Jané-Llopis, E., Manthey, J., Kaner, E., & Rehm, J. (2019). Immediate impact of minimum unit pricing on alcohol purchases in Scotland: Controlled interrupted time series analysis for 2015-18. Bmj, 366.

Palepu, A., Gadermann, A., Hubley, A. M., Farrell, S., Gogosis, E., Aubry, T., & Hwang, S. W. (2013). Substance use and access to health care and addiction treatment among homeless and vulnerably housed persons in three Canadian cities. PloS One, 8(10), e75133.

Pauly, B., Brown, M., Evans, J., Gray, E., Schiff, R., Ivsins, A., Krysowaty, B., Vallance, K., & Stockwell, T. (2019). “There is a Place”: Impacts of managed alcohol programs for people experiencing severe alcohol dependence and homelessness. Harm Reduction Journal, 16(1), 1–14.

Pauly, B., King, V., Smith, A., Tranquilli-Doherty, S., Wishart, M., Vallance, K., Stockwell, T., & Sutherland, C. (2021). Breaking the cycle of survival drinking: Insights from a non-residential, peer-initiated and peer-run managed alcohol program. Drugs: Education, Prevention and Policy, 28(2), 172–180.

Rehm, J., & Probst, C. (2018). What about drinking is associated with shorter life in poorer people? PLoS Medicine, 15(1), e1002477.

Saunders, J. B., Aasland, O. G., Babor, T. F., De la Fuente, J. R., & Grant, M. (1993). Development of the alcohol use disorders identification test (AUDIT): WHO collaborative project on early detection of persons with harmful alcohol consumption-II. Addiction, 88(6), 791–804.

Schofield, L., Walsh, D., Bendel, N., & Piroddi, R. (2021). Excess mortality in Glasgow: Further evidence of ‘political effects’ on population health. Public Health, 201, 61–68.

Scottish Government. (2021, January 11). Glasgow Deprivation Map. Scotland’s Data on a Map. https://datamap-scotland.co.uk/2021/01/glasgow-deprivation-map/

Secombe, P., Campbell, L., Brown, A., Bailey, M., & Pilcher, D. (2021). Hazardous and harmful alcohol use in the Northern Territory, Australia: The impact of alcohol policy on critical care admissions using an extended sampling period. Addiction, 116(10), 2653–2662.

Stockwell, T., Churchill, S., Sherk, A., Sorge, J., & Gruenewald, P. (2020). How many alcohol-attributable deaths and hospital admissions could be prevented by alternative pricing and taxation policies? Modelling impacts on alcohol consumption, revenues and related harms in Canada. Chronic Diseases and Injuries in Canada, 40(5/6).

Stockwell, T., Zhao, J., Giesbrecht, N., Macdonald, S., Thomas, G., & Wettlaufer, A. (2012). The raising of minimum alcohol prices in Saskatchewan, Canada: Impacts on consumption and implications for public health. American Journal of Public Health, 102(12), e103–e110.

Taylor, N., Miller, P., Coomber, K., Livingston, M., Scott, D., Buykx, P., & Chikritzhs, T. (2021). The impact of a minimum unit price on wholesale alcohol supply trends in the Northern Territory, Australia. Australian and New Zealand Journal of Public Health, 45(1), 26–33.

Watts, B., Bramley, G., Fitzpatrick, S., Pawson, H., & Young, G. (2021). The homelessness monitor: Scotland 2021. London: Crisis. https://www.crisis.org.uk/media/245880/the-homelessness-monitor-scotland-2021.pdf

Westenberg, J. N., Kamel, M. M., Addorisio, S., Abusamak, M., Wong, J. S. H., Outadi, A., Jang, K. L., & Krausz, R. M. (2021). Non-beverage alcohol consumption among individuals experiencing chronic homelessness in Edmonton, Canada: A cross-sectional study. Harm Reduction Journal, 18(1), 108. https://doi.org/10.1186/s12954-021-00555-8

World Health Organization. Regional Office for Europe. (2019). Alcohol policy impact case study: The effects of alcohol control measures on mortality and life expectancy in the Russian Federation. World Health Organization. Regional Office for Europe. https://apps.who.int/iris/handle/10665/328167

World Health Organization. Regional Office for Europe. (2021). Making the WHO European Region SAFER: Developments in alcohol control policies, 2010–2019. World Health Organization. Regional Office for Europe. https://doi.org/10.37426/9789289055048

World Health Organization. Regional Office for Europe. (2022). No place for cheap alcohol: The potential value of minimum pricing for protecting lives. World Health Organization. Regional Office for Europe. https://apps.who.int/iris/handle/10665/356597

Zeitler, M., Williamson, A. E., Budd, J., Spencer, R., Queen, A., & Lowrie, R. (2020). Comparing the impact of primary care practice design in two inner city UK homelessness services. Journal of Primary Care & Community Health, 11, 2150132720910568.

Zhao, J., & Stockwell, T. (2017). The impacts of minimum alcohol pricing on alcohol attributable morbidity in regions of British Colombia, Canada with low, medium and high mean family income. Addiction, 112(11), 1942–1951.

Zhao, J., Stockwell, T., Martin, G., Macdonald, S., Vallance, K., Treno, A., Ponicki, W. R., Tu, A., & Buxton, J. (2013). The relationship between minimum alcohol prices, outlet densities and alcohol-attributable deaths in B ritish C olumbia, 2002–09. Addiction, 108(6), 1059–1069.

